# Genomic epidemiology of SARS-CoV-2 divulge B.1, B.1.36, and B.1.1.7 as the most dominant lineages in first, second, and third wave of SARS-CoV-2 infections in Pakistan

**DOI:** 10.1101/2021.07.28.21261233

**Authors:** Atia Basheer, Imran Zahoor

## Abstract

The present study aims to investigate the genomic variability and epidemiology of SARS-CoV-2 in Pakistan along with their role in the spread and severity of infection during the three waves of COVID-19. A total of 453 genomic sequences of Pakistani SARS-CoV-2 were retrieved from GISAID and subjected to MAFFT-based alignment and QC check which resulted in removal of 53 samples. The remaining 400 samples were subjected to Pangolin-based genomic lineage identification. And to infer our SARS-CoV-2 time-scaled and divergence phylogenetic trees, 3,804 selected global reference sequences plus 400 Pakistani samples were used for the Nextstrain analysis with Wuhan/Hu-1/2019, as reference genome. Finally, maximum likelihood based phylogenetic tree was built by using the Nextstrain & coverage map was created by employing Nextclade. And by using the amino acid subsitutions the maximum likelihood phylogenetic trees were developed for each wave, separately. Our results reveal the circulation of 29 lineages, belonging to following 7 clades G, GH, GR, GRY, L, O, & S in the three waves. From first wave, 16 genomic lineages of SARS-CoV-2 were identified with B.1(24.7%), B.1.36(18.8%), & B.1.471(18.8%) as the most prevalent lineages respectively. The second wave data showed 18 lineages, 10 of which were overlapping with the first wave suggesting that those variants could not be contained during the first wave. In this wave, a new lineage, AE.4, was reported from Pakistan for the very first time in the world. However, B.1.36 (17.8%), B.1.36.31 (11.9%), B.1.1.7 (8.5%) & B.1.1.1 (5.9%) were the major lineages in second wave. Third wave data showed the presence of 9 lineages with Alpha/B.1.1.7 (72.7%), Beta/B.1.351 (12.99%), & Delta/B.1.617.2 (10.39%) as the most predominant variants. It is suggested that these VOCs should be contained at the earliest in order to prevent any devastating outbreak of SARS-CoV-2 in the country.

## Introduction

Since the initial reports of coronavirus disease 2019 (COVID-19) outbreak in Wuhan, China on 30 December 2019 [1, 2], SARS-CoV-2 has been spreading worldwide and, as of 15 July 2021, there have been 188.13 million confirmed infections and 4.06 million deaths [3]. This was the reason that SARS-CoV-2 epidemic was declared a global pandemic by World Health Organization (WHO) on 11 March 2020 [4]. SARS-CoV-2 is a single-stranded positive-sense RNA virus belonging to the genus *Betacoronavirus*, and subgenus *Sarbecovirus*. The genome size of SARS-CoV-2 is approximately 30kb and its genomic structure has followed the characteristics of known genes of the coronavirus [5]. Albeit, RNA viruses are highly prone to mutations but Coronaviruses (CoVs) have an outstanding and distinctive feature of intrinsic proofreading mechanism [6]. However, CoVs encode a protein called nonstructural protein 14 (nsp14) that possesses a 3’ to 5’ exonuclease (ExoN) activity which is necessary for replication fidelity and proofreading activity [7]. Owing to the presence of large and complex genome, this proofreading mechanism is considered critical for maintaining normal functioning and fitness of CoVs. And, perhaps, this is the reason that mutation rate in SARS-CoV-2 is 10-fold lesser compared with other RNA viruses. However, even then geneticists had reported a rate of 33 mutations/year in SARS-CoV-2 genome and now scientists are using these mutations to categorize different variants of this virus into different clades, lineages, and sublineages [4]. To date the SARS-CoV-2 has been divided in more than 81 lineages, on the basis of variations in its genome, which vary significantly in their transmissibility and virulence [8]. All the existing lineages including variants of concerns are the descendants of two ancestral lineages, A and B, discovered from China in the start of the pandemic. However, European Center for Disease Prevention and Control (ECDC) has demonstrated five variants of concerns (VOCs) due to their high transmissibility, pathogenicity, and effects on the vaccine efficacy and that’s why they are known as VOCs. Alpha variant (B.1.1.7) also known as Alpha VOC, was first detected in UK in September 2020 [9]. This variant is highly variable and contain more than a dozen mutations compared with wild-type lineages. It had also been reported to have 50-70% high transmissibility [10], greater severity of the disease [11, 12], and effects on the efficacy of the vaccine compared with wild-type lineage. Beta variant of concern, B.1.351, also known as 20H/501Y.V2 was identified from South Africa. It shares several mutations with B.1.1.7 and has the following major amino acid mutations, K417N, E484K, N501Y, D614G, & A701V in its spike protein. This lineage is also reported to have increased transmissibility and even have the advantage of escaping from immunity due to the presence of E484K mutation in its spike protein [13]. Likewise, B.1.617.2 lineage (Delta variant), also known as 20A/S:478K, was identified in India at the end of 2020 and had been reported to have the following major spike mutations, L452R, T478K, D614G, & P681R, which are involved in enhancing its transmissibility and risk of hospitalization compared with B.1.1.7 [14-16]. Lineage B.1.617.2, also known as Delta variant of concern was detected from India in December 2020 with following mutation of interest L452R, T478K, D614G, P681R in its spike protein. Likewise, P.1 lineage (Gamma), also known as 20J/501Y.V3, was identified from Brazil [17]. It has K417T, E484K, N501Y, D614G, and H655Y as the most noteworthy amino acid mutations which have impact on its transmissibility and immunity [18]. B.1.427 and B.1.429 lineages (Epsilon), also collectively known as 20C/S452R [19] are two more variants of concern which are based on several spike protein mutations, including L452R, which is associated with increased cell entry and reduced susceptibility to neutralization by convalescent and vaccine recipient plasma in vitro [20].

In Pakistan, first two cases of COVID-19 were confirmed on February 26, 2020 and by June 17, 2020 each region of Pakistan has reported at least one confirmed case of Coronavirus disease. Pakistan has experienced three different waves of COVID-19, to date. The first nationwide COVID-19 wave started in late May 2020 and reached its peak in mid-June, when the number of newly confirmed cases and the number of daily deaths reached at its peak, and then in mid-July, it ended abruptly. The second wave of COVID-19 started in early November 2020 but the intensity of this wave was relatively low, and it mainly affected the southern part of Sindh. The country’s third wave began in mid-March 2021 and it mainly affected the Punjab and Khyber Pakhtunkhwa provinces. This wave peaked in late April 2021, and since then, number of new cases and daily deaths have been decreasing. In Pakistan, 978,662 individuals have been infected with SARS-CoV-2 and 22,642 of them have died until 15 June 2021. The first three cases of B.1.1.7 (UK-VOC) were reported on 26 February 2021, and perhaps this was the reason that the third wave of COVID-19 was much severe compared with first two waves. Hence, it is hypothesized that multiple variants of SARS-CoV-2 are present in Pakistan which are perhaps the reason for the repeated occurrence of COVID-19 epidemics in the country. However, the scarcity of genomics and epidemiologic studies on the transmission, spread, and distribution of different variants of virus in Pakistan has further hampered the control efforts in the country. Thus, the aims of the current study are to identify the lineages along with their origin, transmission, and prevalence in the country and the estimation of genetic relatedness among the variants, present in Pakistan and world, through phylogenetic clustering by using the whole genome sequence data.

## Results

The data about the health status of COVID-19 patients in different geographical regions of Pakistan, taken from official website of Pakistan (www.covid.gov.pk), is presented in Table 1 and Fig 1.

**Table 1.** The data of COVID-19 taken from official covid website (http://www.covid.gov.pk) of Pakistan for a period spanning from 3 January 2020 to 16 June 2021.

| Areas | Diseased | Hospitalized | Deceased | Recovered |
| --- | --- | --- | --- | --- |
| Azad Jammu & Kashmir (AJK) | 19,868 | 447 | 571 | 18850 |
| Baluchistan | 26,446 | 794 | 300 | 25372 |
| Gilgit-Baltistan | 5,759 | 105 | 108 | 5546 |
| Islamabad | 82,278 | 1216 | 773 | 80289 |
| Khyber Pakhtoon Khawah (KPK) | 136,663 | 2958 | 4252 | 129453 |
| Punjab | 344,641 | 10462 | 10603 | 323576 |
| Sindh | 330,552 | 19827 | 5306 | 305419 |

**Fig. 1.**
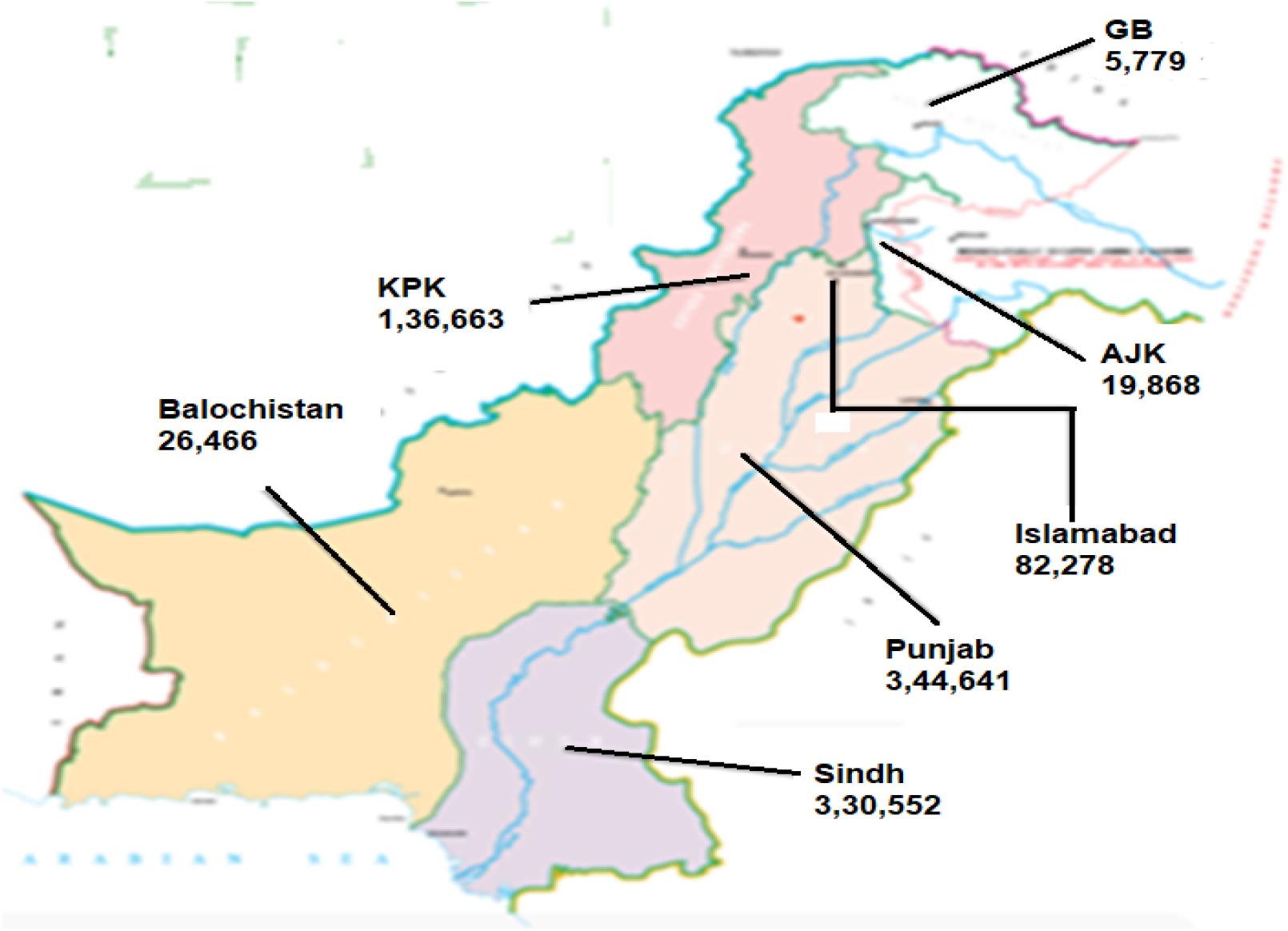
Number of the total cases of COVID-19 in different provinces/territories of Pakistan up to 16 June 2021. COVID cases of Islamabad, Capital of Pakistan, are also shown here. This map is adopted from https://covid.gov.pk/.

It is observed that maximum COVID-19 cases were detected in Punjab (344,641) followed by Sindh (330,552), KPK (136,663), Baluchistan (26,446), AJK (19,868), Capital city of Islamabad (82,278) and Gilgit-Baltistan (5,759) region of Pakistan (Fig. 1). During this duration, 21,913 deaths were recorded in Pakistan, out of which the highest numbers (10,603) were observed in Punjab province.

### Major waves of SARS-CoV-2 infection in Pakistan

Pakistan has observed three major waves of SARS-CoV-2 infection. The nation’s first wave of SARS-CoV-2 began in late May 2020, peaked in mid-June and ended in mid-July. In this wave, Pakistan experienced the highest number of cases and mortalities during its peak month. However, this wave passed away very rapidly and new cases and mortalities were dropped suddenly after the peak. Second surge of COVID-19 started from end of October 2020 to mid-February 2021. This wave was low in its intensity, mainly affecting the southern parts of Sindh province, and peaked in mid-December 2020. The country’s third wave began in mid-March 2021, when daily new confirmed cases and mortalities began to rise steeply. The third wave mainly affected the provinces of Punjab and Khyber Pakhtunkhwa. This wave peaked in late April 2021, and since then daily new cases, and new mortalities have been falling. The complete pictures of number of SARS-CoV-2 positive cases and mortalities from February 2020 to May 2021 is shown in Fig. 2 and 3.

**Fig 2.**
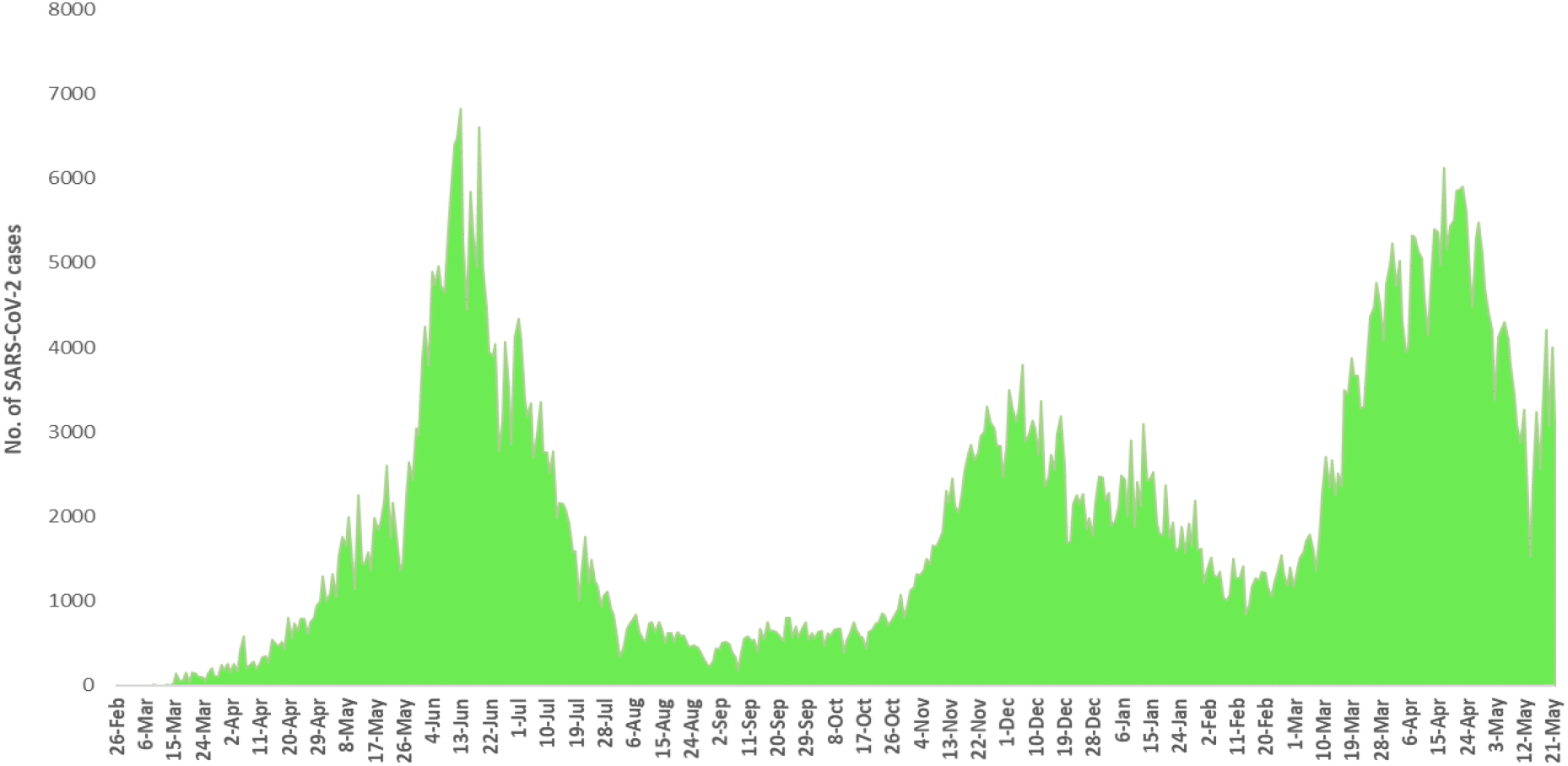
Month-wise details of SARS-CoV-2 cases starting from February 2020 to May 2021

**Fig 3.**
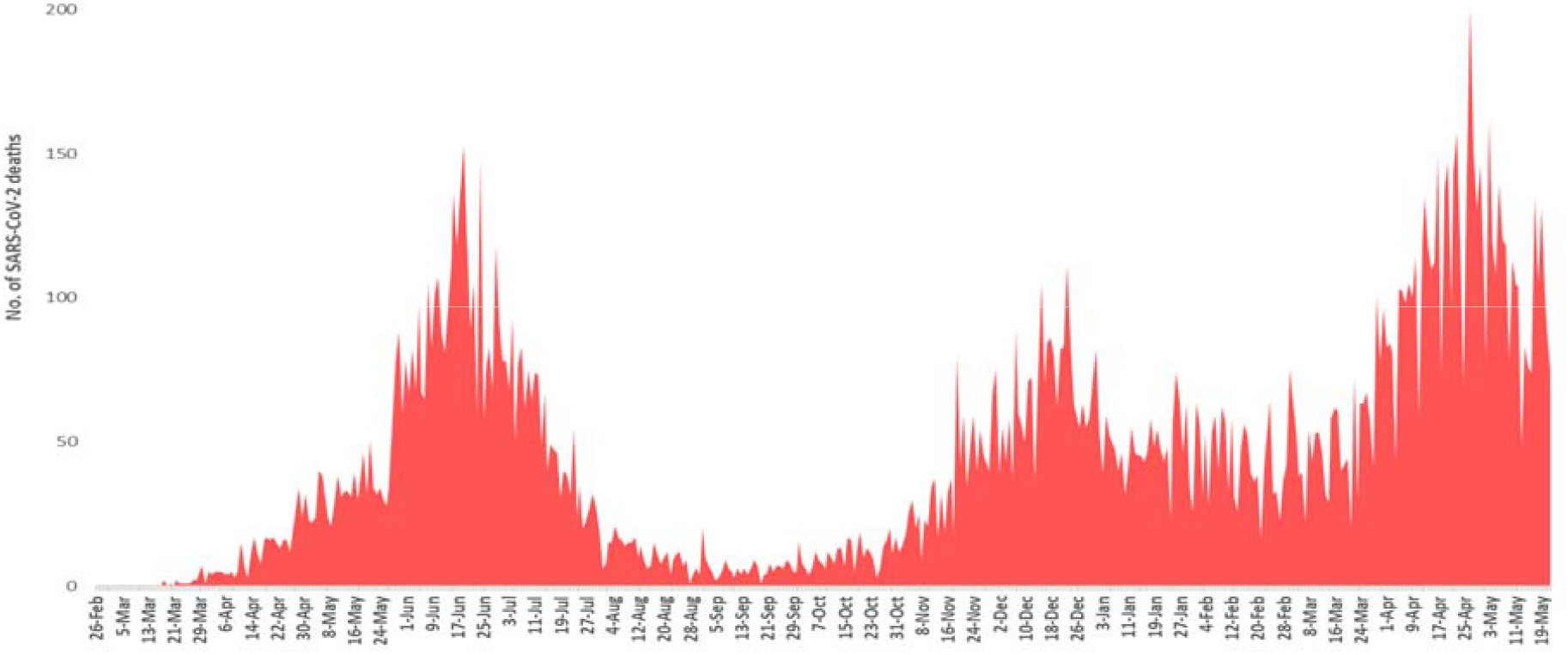
Month-wise details of deaths caused by SARS-CoV-2 starting from February 2020 to May 2021

### Distribution of Genomic Lineage of SARS-CoV-2 in Pakistan

In the current study, a total of 453 SARS-CoV-2 genomes sequences downloaded from GISAID on 10 July 2021 were used. In Pakistan, 29 lineages were observed by using Phylogenetic Assignment of Named Global Outbreak Lineage (http://www.pangolin.cog-uk.io). Out of total, 53 samples were failed. The overall lineage distribution highlighted the occurrence of B.1.1.7(n=178), B.1.36(n=39), B.1.351(n=30), B.1(n=25), B.1.617.2(n=24), B.1.471(n=22), B.1.36.31(n=18), B.1.1.1(n=10)), B.1.1(n=9), A(n=9), C.23(n=6), B(n=5), B.1.36.17(n=1), B.1.36.34(n=5), B.6(n=3), B.1.499(n=2), B.1.1.448(n=2), AE.4(n=1), B.1.260(n=1), B.1.36.24(n=1), B.1.36.8(n=1), B.1.468(n=1), B.1.523(n=1), B.1.562(n=1), B.1.1.25(n=1), B.1.1.372(n=1), B.1.1.413(n=1), B.4(n=1) and B.6.6(n=1) as mentioned in Fig. 4 and Table 2.

**Fig. 4.**
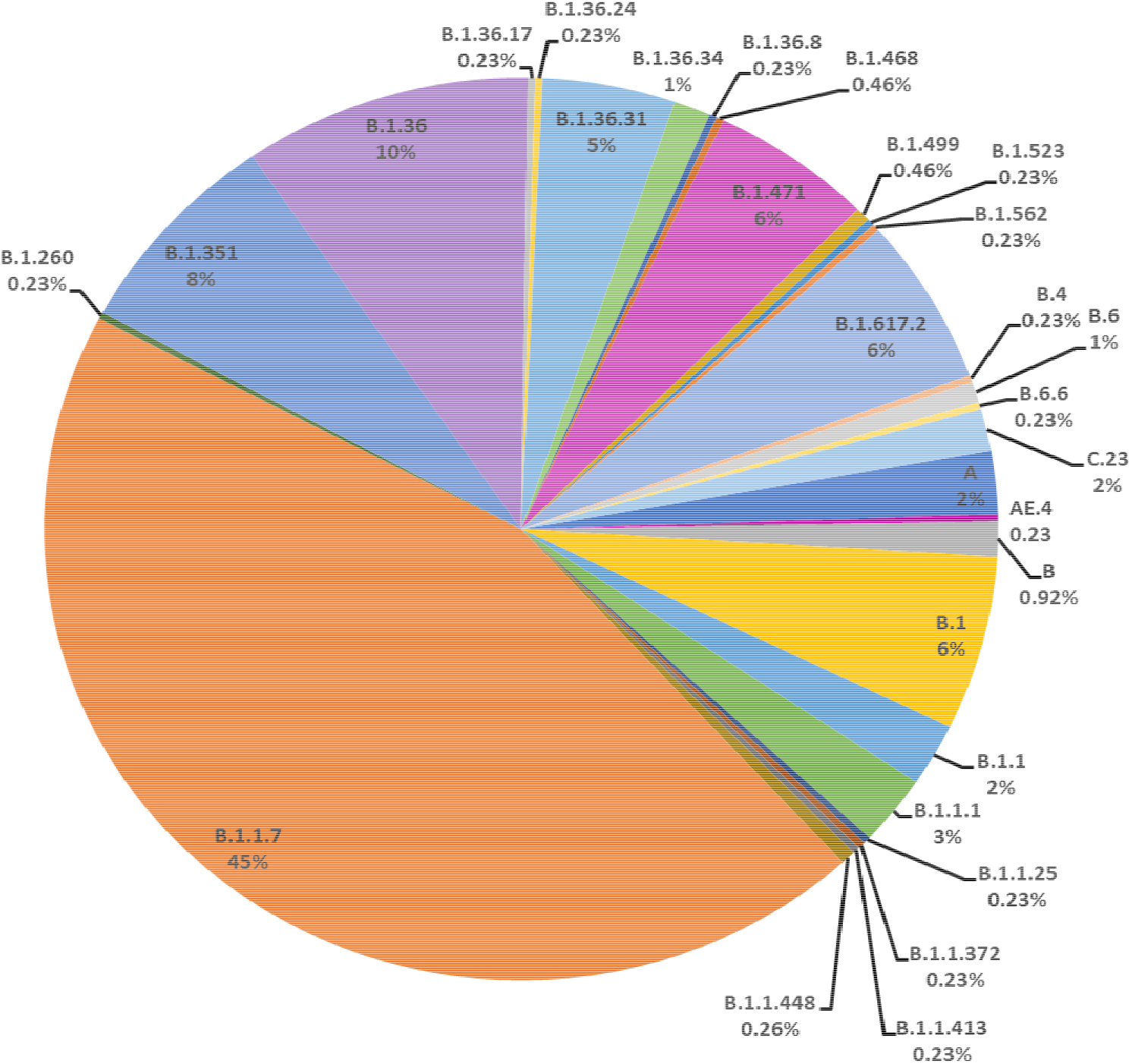
All the lineages observed from Pakistan sequences downloaded on 10 July 2021.

**Table 2.**
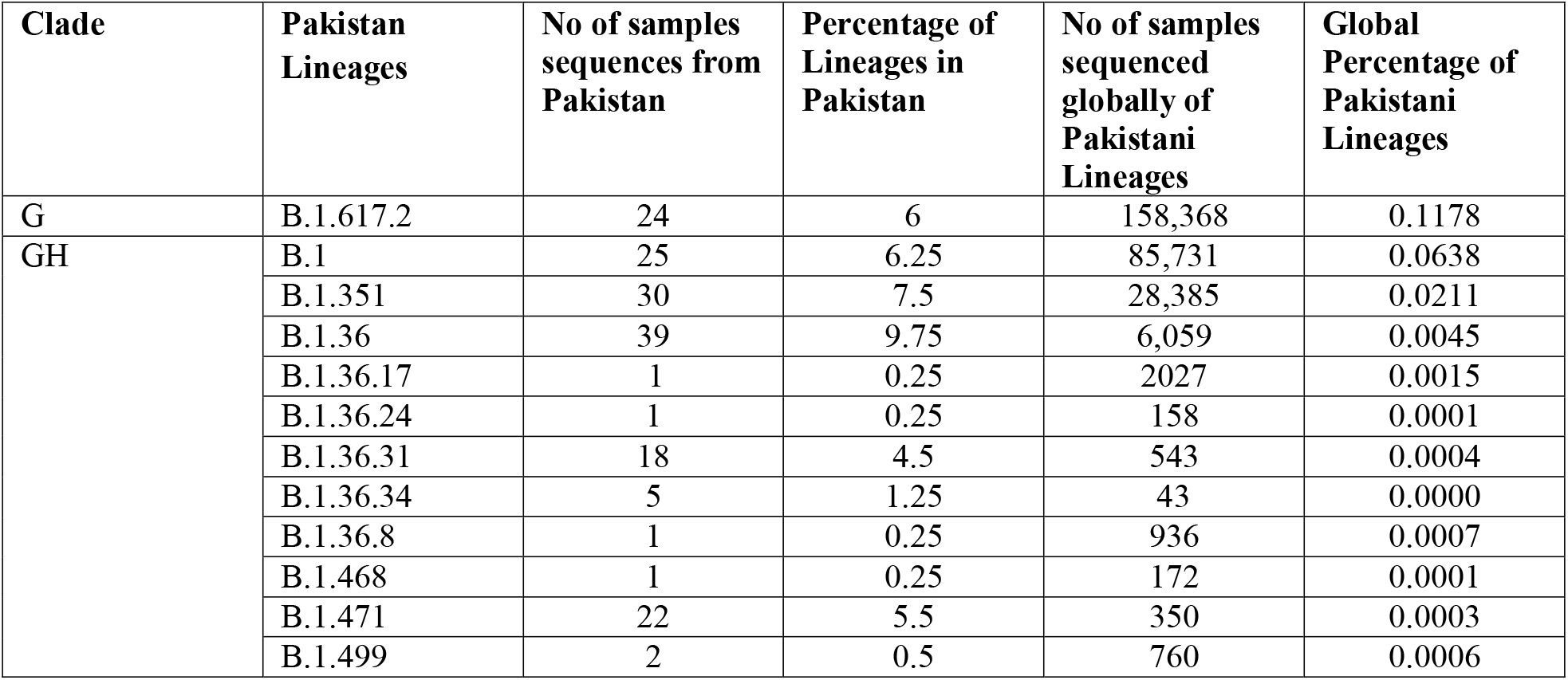
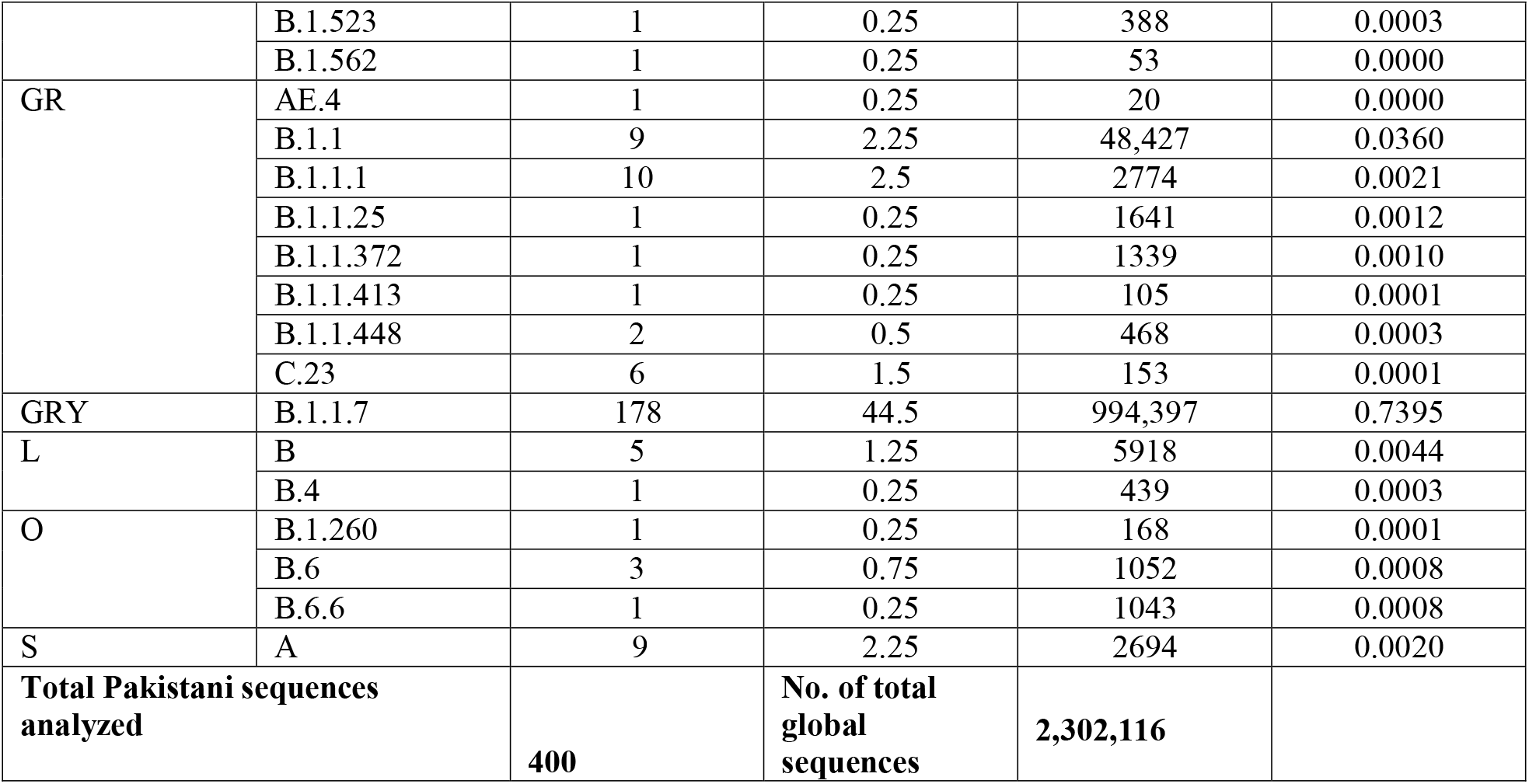
List of 29 lineages of SARS-CoV-2 observed in Pakistan along with their local and global percentages.

### Distribution of GISAID clades of SARS-CoV-2 in Paksitan

The distribution of GISAID clades of SARS-CoV-2 genomes from Pakistani & global dataset, downloaded on 10 July 2021, is presented in Table 2 & Fig 5. The genomic sequences of Pakistani SARS-CoV-2 samples indicate the prevalence of 7 clades with following order of prevalence GRY (44.5%), GH (36.75%), GR (7.75%), G (6%), S (2.25%), L (1.50%), & O (1.25%).

**Fig 5.**
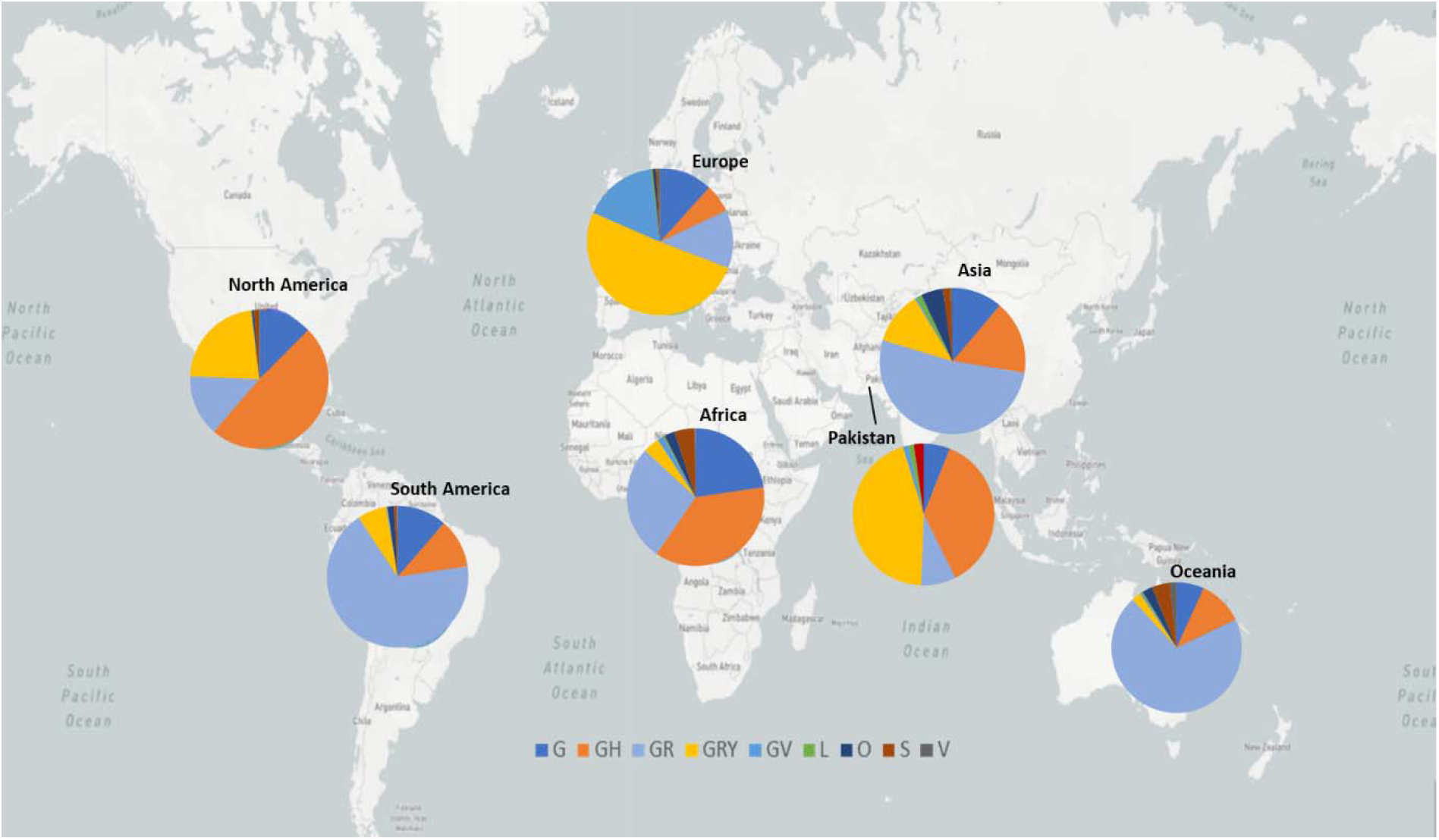
GISAID clades of SARS-COV-2 genome from global and Paksitani datasets. The map in the background is adopted from https://nextstrain.org/sars-cov-2/ [21].

### Occurrence of Variants of concerns in Pakistan

Our data also revealed the presence of the three VOCs including Alpha (B.1.17), Beta (1. 351) and Delta (B.1.617.2) variants in Pakistan, with the following order of genomic prevalence 44.5%, 7.5% and 6% respectively.

### Phylogenetic Analysis

Maximum Likelihood Phylogenetic tree of 400 whole genome sequences of Pakistani SARS-CoV-2 was developed as per definitions of the PANGOLIN lineage and GISAID clades (Fig 7). The overall clades and lineages distribution highlighted the dominant occurrence of GRY [B.1.1.7], GH [B.1, B.1.351, B.1.36, B.1.36.17, B.1.36.24, B.1.36.31, B.1.36.34, B.1.36.8, B.1.468, B.1.471, B.1.499, B.1.523, B.1.562], GR [AE.4, B.1.1, B.1.1.1, B.1.1.25, B.1.1.372, B.1.1.413, B.1.1.448, C.23], G [B.1.617.2], S [A], L [B, B.4] and O [B.1.260, B.6, B.6.6] clades respectively. Maximum likelihood time resolved phylogeny tree of Pakistani and global samples is presented in Fig 8. The genomic map of SARS-CoV-2 of Pakistani samples depicting the mutational profiles is in given in Fig 6.

**Fig 6.**
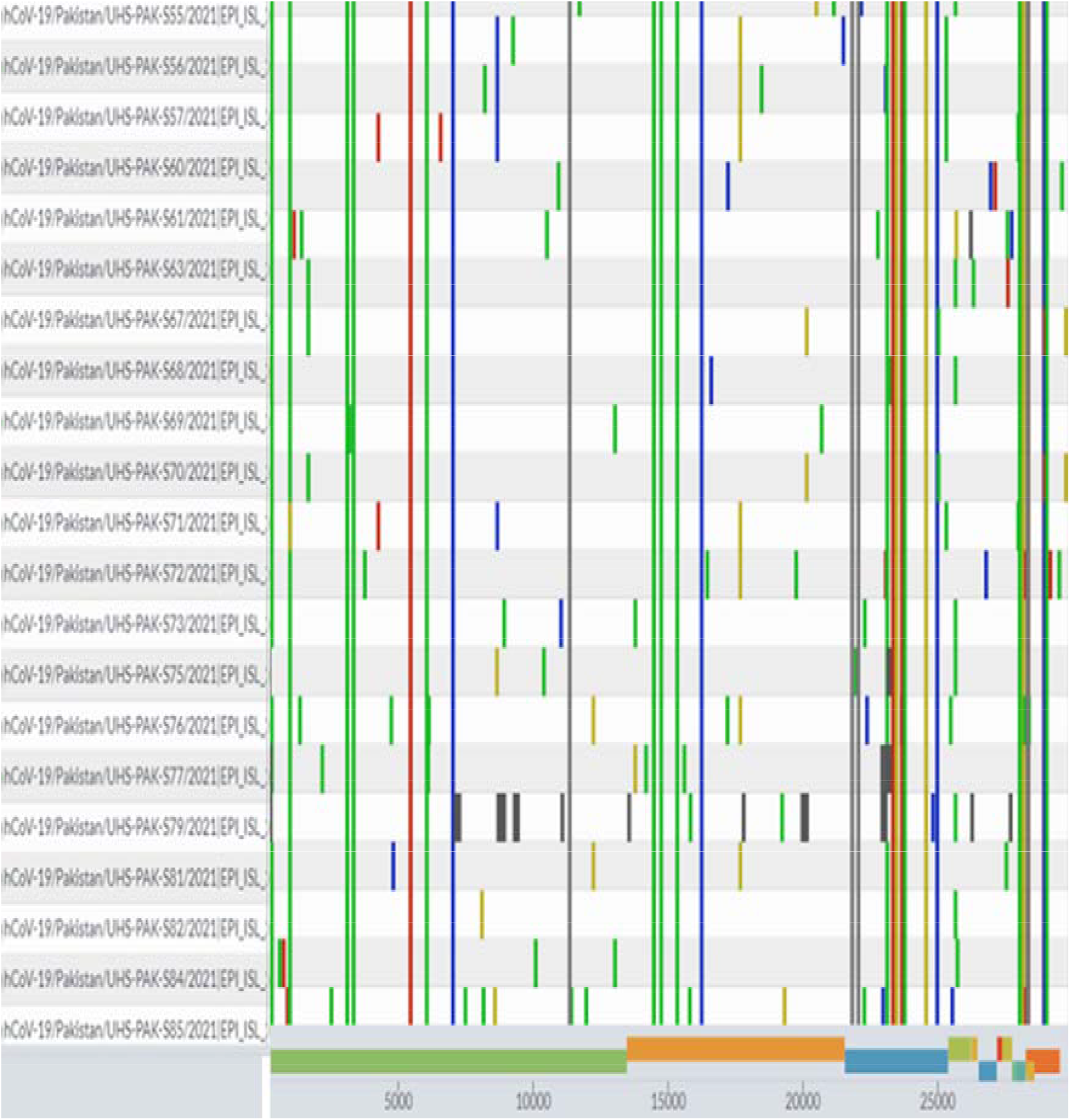
Genomic map of SARS-CoV-2 genomes of Pakistan. The reference genome in this coverage map is Wuhan/Hu-1/2019.

**Fig 7.**
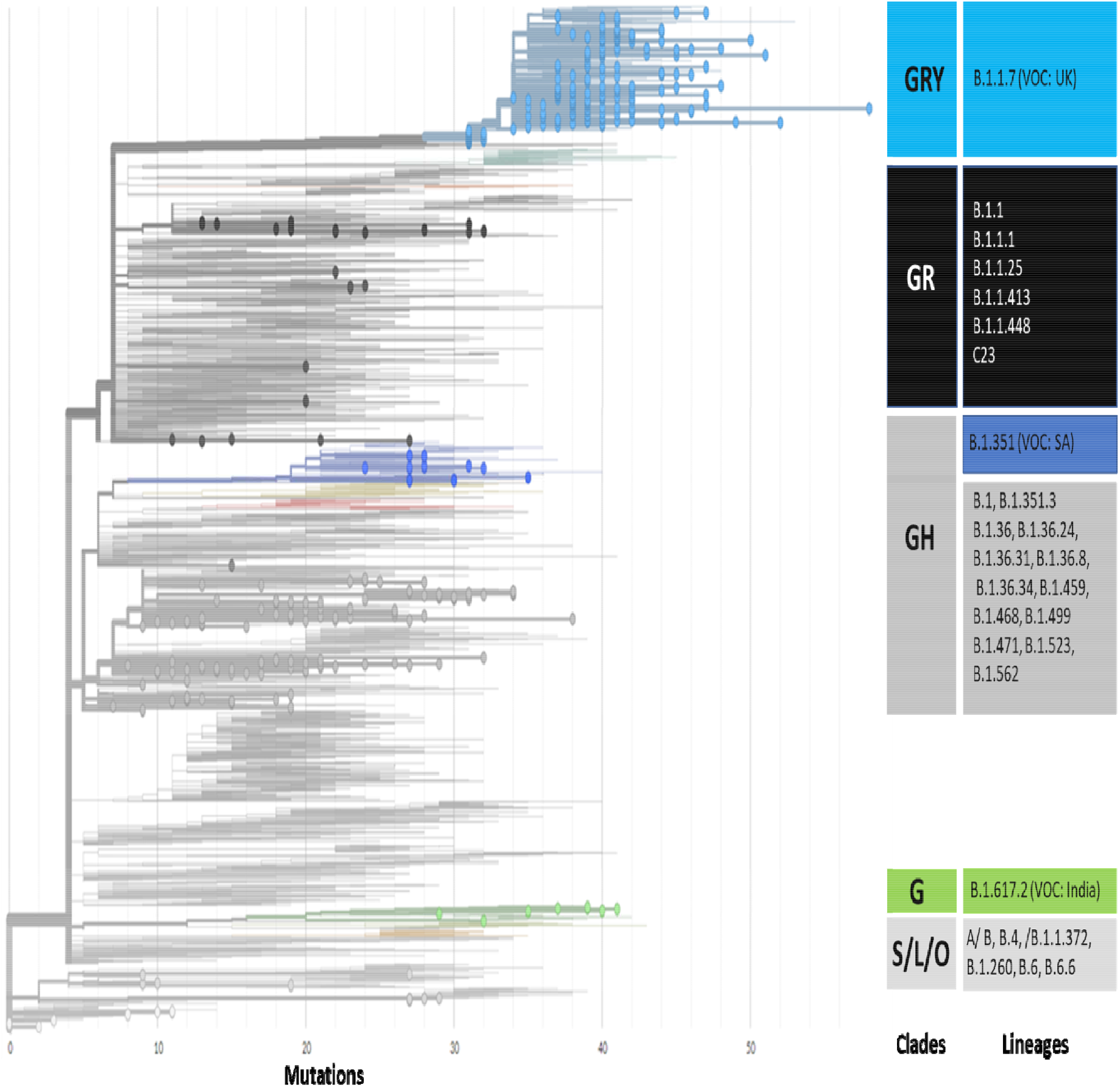
Maximum likelihood phylogenetic tree of Pakistani samples showing 7 clades and 29 lineages. The global SARS-CoV-2 genomes depicting the highest mutations ranging from 30 to 50 in Pakistani samples under the lineage of B.1.1.7, also known as UK variants of concern (UK-VOC).

**Fig 8.**
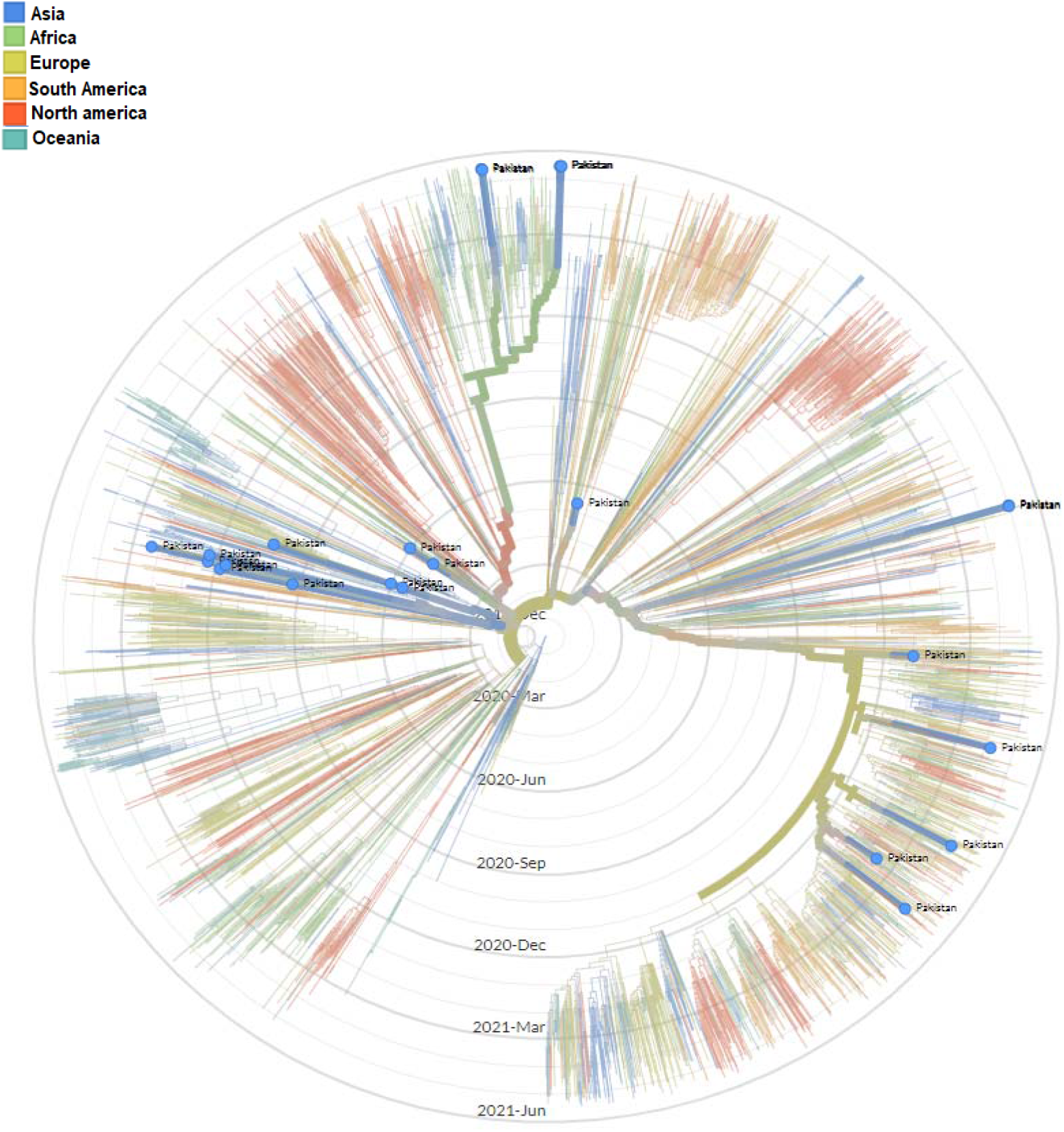
Maximum Likelihood time resolved phylogenetic tree of global samples representing the emergence of SARS-CoV-2 in Pakistan.

### Wave-wise phylogenetic trees of SARS-CoV-2

Maximum likelihood phylogenetic tree of first, second and third wave are presented in Fig. 9, 10 and 11. The tree of the first wave showed 16 lineages of 5 clades, with GH as major clade in this wave (Fig. 9). During second wave 19 lineages belonging to four clades were observed and GH was again major clade in this wave (Fig. 10). In third waves, 9 lineages representing 5 clades were observed (Fig. 11), and GRY was the major clade in this wave (Fig. 12).

**Fig 9.**
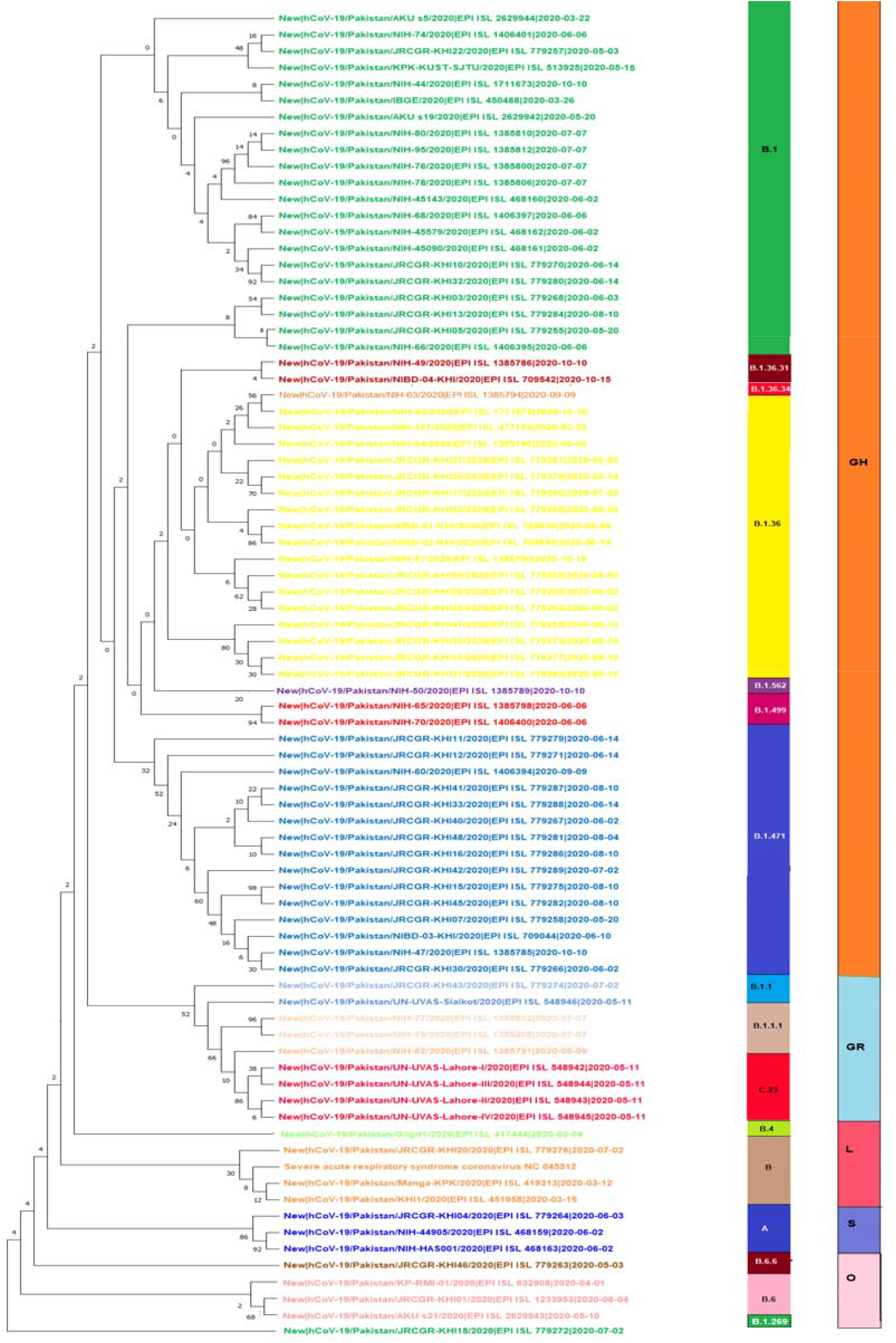
Maximum Likelihood phylogenetic tree of SARS-CoV-2 genome representing 16 lineages of 5 clades in first wave.

**Fig 10.**
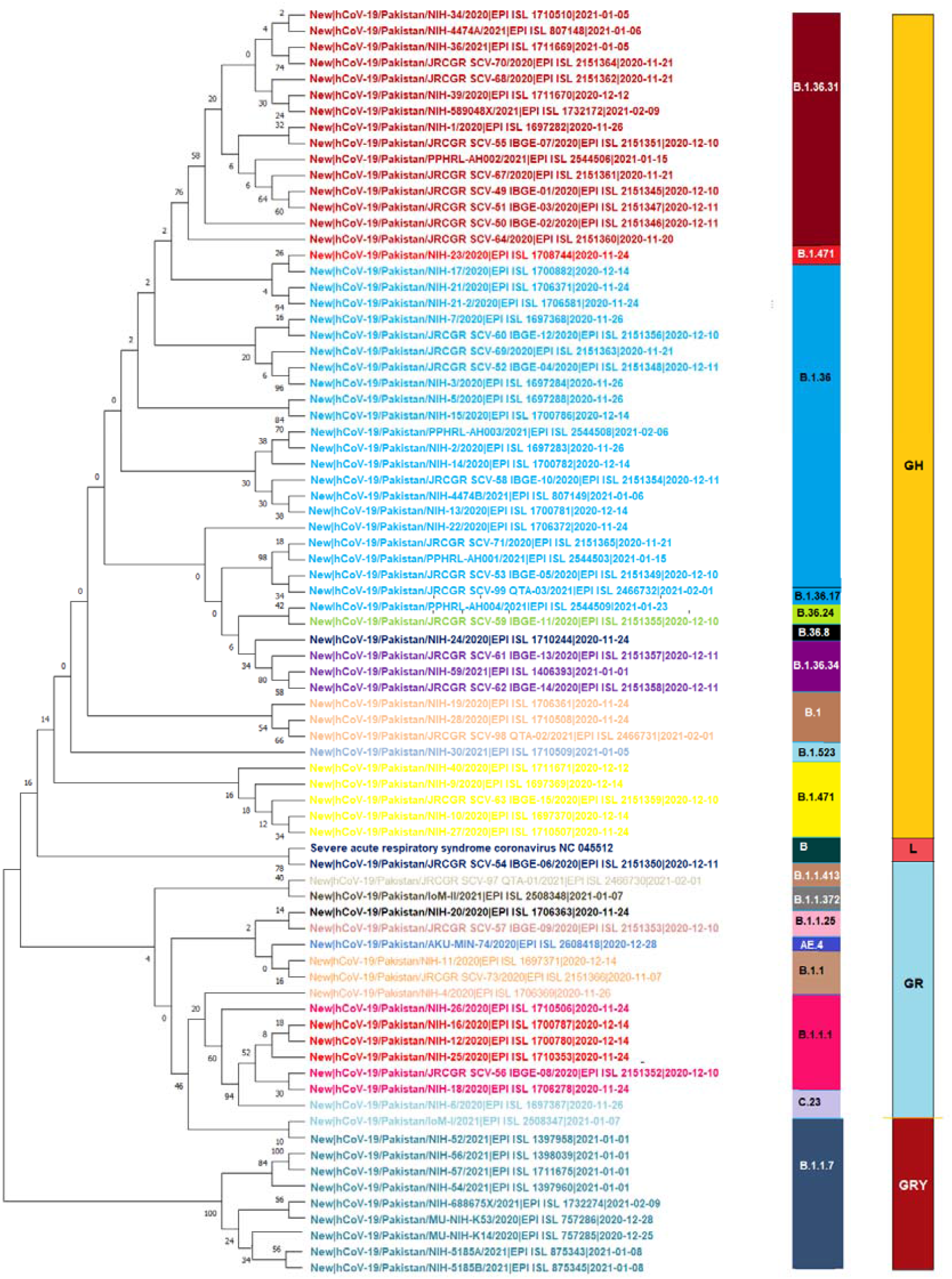
Maximum Likelihood phylogenetic tree of SARS-CoV2 genome representing 19 lineages and 4 clades in second wave.

**Fig 11.**
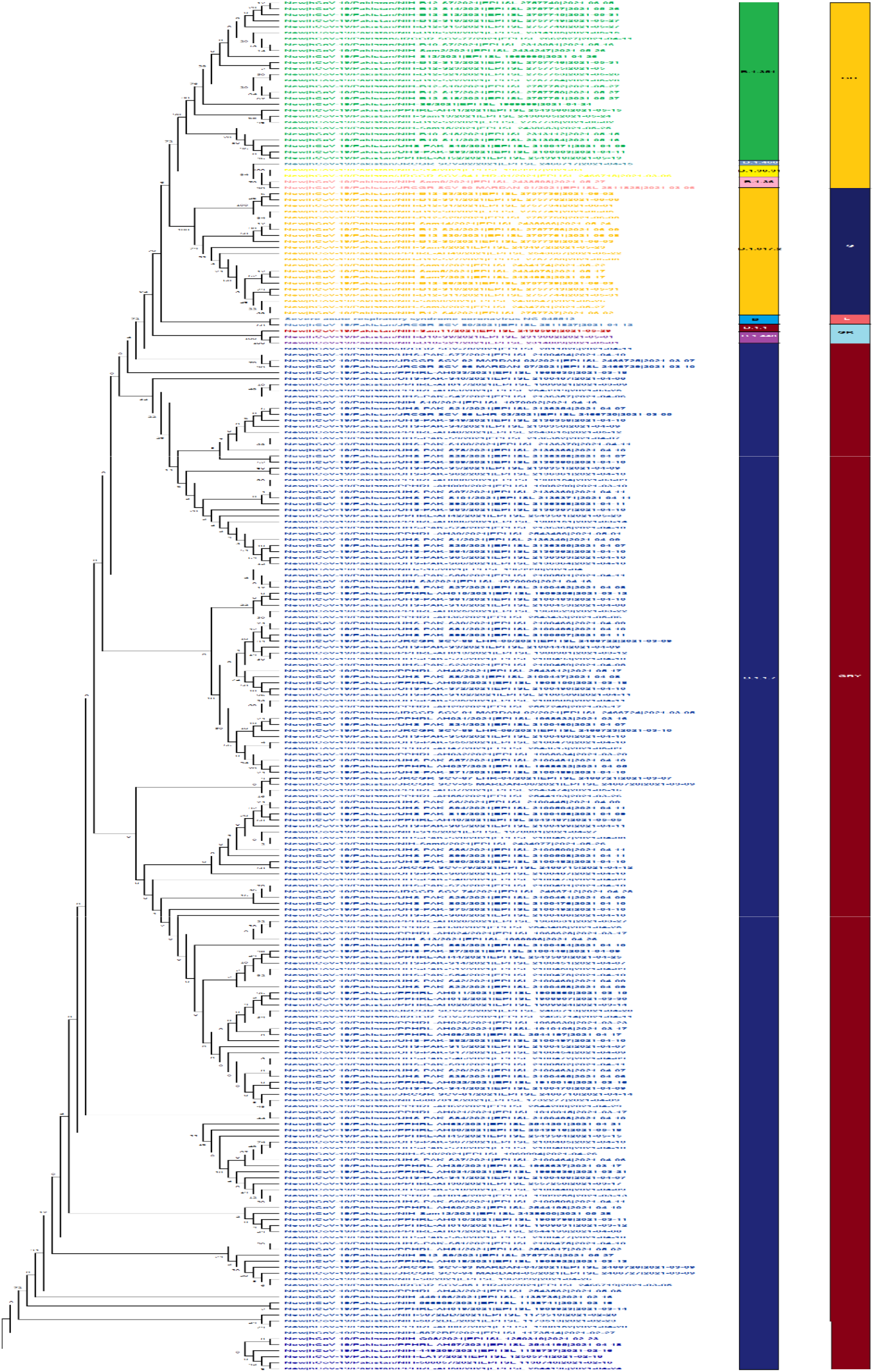
Maximum Likelihood phylogenetic tree of SARS-CoV-2 genome representing 9 lineages and 5 clades in third wave.

**Fig 12.**
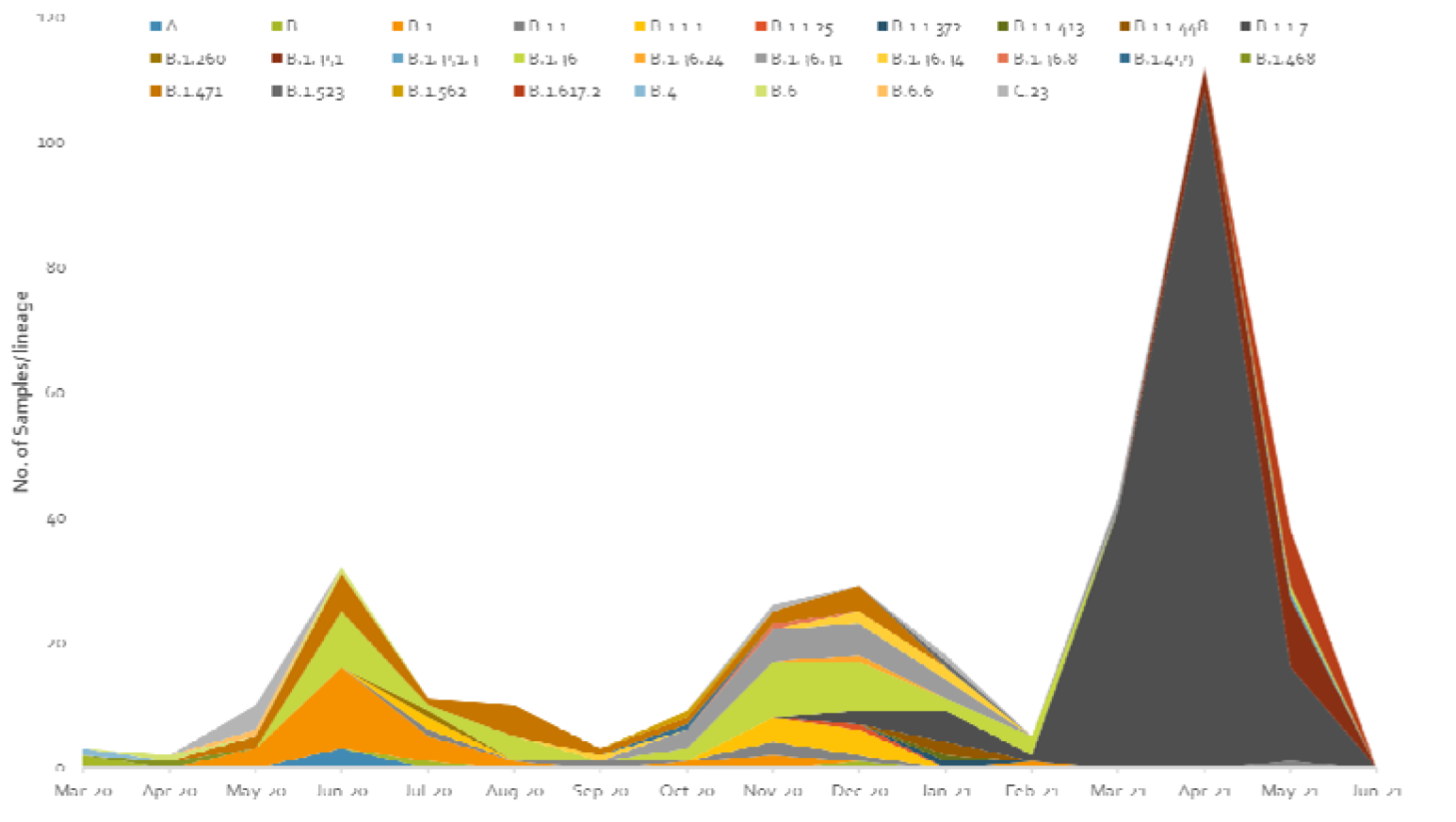
Pictorial view of genomic-data-based incidence of different lineages of SARS-CoV-2 observed during the three waves of COVID-19 in Pakistan

## Discussion

First two cases of COVID-19 were reported in Pakistan in February 2020 and since then country has observed 978,662 infections and 22,642 deaths till 15 July 2021. However, the whole genome sequences of first three SARS-CoV-2 samples, collected in March, revealed B and B.4 lineages. The lineage B.4 was reported to had major role in the early pandemic in Iran [22]. This supports the hypothesis of an early transmission of virus to Pakistan most likely from Iran and China, and this is also consistent with statement of Pakistan’s health ministry officials who confirmed first two cases of COVID-19 on 26^th^ of February from patients travelled back to Pakistan from Iran. It can be further confirmed by the fact that up to the first week of April 2020, 75% of the corona positive patients had the history of traveling back to Pakistan from Iran [23]. The occurrence of severe outbreaks of COVID-19 in two neighboring countries and declaration of the disease as a pandemic by WHO prompted the Pakistan government to close the border with China and put very strict screening methods at the Pak-Iran border [24, 25]. And by following the guidelines of Federal government devised *“The National Action Plan for The Corona Virus Disease (COVID-19) Pakistan”*, the quarantine centers were established in few big cities including Islamabad [26, 27] and Pak-Iran border to help identify positive cases and quarantine pilgrims returning back to Pakistan after spending time in Iran [28]. However, even with all of these efforts, major lapses existed at every step mainly the uneven implementation of immigrant policies to deal with the influx of people from the outside of the country [29], the lack of facilities, poor infrastructure, and apathy & indifference among the peoples to follow government devised Corona SOPs led to the spread of COVID-19 throughout the country [30].

### First wave of SARS-CoV-2

Pakistan experienced three major waves of SARS-CoV-2 infection. First wave mainly covered three months starting from May 2020 to July 2020 (Fig 2 & 3). From this wave, whole genome sequences of 86 samples of SARS-CoV-2 were analyzed. These samples represented 16 lineages belonging to 5 clades (Fig 9) including A(9.3%), B(4.6%), B.1(24.4%), B.1.1(2.3%), B.1.1.1(3.4%), B.1.260(1.16%), B.1.36(18.6%), B.1.36.31(2.32%), B.1.36.34(1.2%), B.1.471(18.6%), B.1.499(2.32%), B.1.562(1.2%), B.4(1.2%), B.6(3.4%), B.6.6(1.2%), and C.23(4.65%). Lineage B.1 hit highest score in the first wave in Pakistan and internationally this lineage was reported to have very high incidence in North America (the highest incidence), UK, Germany, and Spain [31, 32]. It is also noteworthy that the highest proportion of Pakistani diaspora is living in these countries. Therefore, it is highly likely that the transmission of this lineage to Pakistan took place through international travelers most probably through the travelling of Pakistani diaspora living in these countries. Though Pakistan suspended all international flights from 21 March to 30 May 2020 but it is highly likely that before the suspension of flight operations [33] many international passengers (positive for these lineage) managed to enter the country. This hypothesis is further augmented from the fact that almost all the sequence samples of this lineage were from big cities including Karachi, Capital city of Islamabad, & Lahore, which have international airports. The lineage for second highest incidence of infection of SARS-CoV-2 in Pakistan was B.1.36. This lineage had the highest incidence of infections in India, Canada, UK and Hong Kong. Likewise, B.1.471 was also the second most prevalent lineage with an incidence of 18.6%. The whole genome sequence data of SARS-CoV-2 from GISAID predict the first incidence of this lineage in Saudi Arabia on 25^th^ March 2020 [34]. After that, this lineage was observed in Pakistan suggesting the direct transmission of this lineage from Saudi Arabia to Pakistan. Likewise, the most likely reason for the transmission of this lineage to Pakistan was again through the international travelers. Another possible explanation for the transmission of these lineages could be the congregation (2,50,000 peoples) of Tablighi Jamaat in Pakistan in which in addition to Pakistan, 3000 delegates from 40 different countries did participate, with significant proportion of delegates from India [35]. This hypothesis could be further confirmed from the timeline of the reports of these lineages in Pakistan as the congregation took place in early March 2020 and these lineages were reported for the first time in May and had the highest incidence in June 2020.

It is also worth mentioning that first 4 cases of C.23 lineage were reported for the first time from Pakistan on 11-05-2020 and 3-4 week later this lineage was also reported from USA. It is also highly likely that this lineage was transmitted to USA from Pakistan through the Pakistani diaspora returning back to USA after visiting their homeland. However only 148 cases of this lineage are reported globally up to 15 July 2021 indicating the lesser transmissibility of this lineage.

### Second wave of SARS-CoV-2

The second spell of SARS-CoV-2 infection in Pakistan ranged from the end of October 2020 to mid of February 2021, however, it was at its peak from mid of November to mid of December 2020 (Fig 2 & 3). For this wave, after QC analysis 35 incomplete sequence of SARS-CoV-2 were removed and remaining 83 samples with whole genome sequences, available on GISAID, were analyzed. According to our dataset, 18 lineages belonging to 4 clades (Fig. 10) were observed in this wave out of which 10 were the same lineages which were already prevailing in the country during the first wave and 8 others were the new lineages. The B.1.36 (initially reported from India, Canada, and UK) which was the second most common (18.6%) lineage during the first wave in Pakistan, had the highest incidence in the second wave as well. And interestingly it was also observed that after the transmission of this lineage to Pakistan it remained prevalent in the country and positive cases of this lineages were observed in almost every month during the first and second wave. The B.1.36.31 (a sub-lineage of B.1.36) which was also present in the first wave had the second highest incidence in second wave. The first case of B.1.36.31 lineage was reported from Australia in the early March 2020 and then two cases of this lineage were reported from England, and 4^th^ case was reported from Pakistan. The incidence of this lineage in the second wave remained 16.9% and out the total 532 genomic sequences, 18 were reported from the second wave in Pakistan (mostly from Karachi).

Likewise, the lineage B.1.471 which also had the second highest (18.6%) incidence during the first wave, remained the third most common lineage in the second wave but its incidence was reduced to 7.23%. It is also noteworthy that only 346 samples of this lineage were observed out of global sequence data available at GISAID, out of which 22 were reported from Pakistan only. And this lineage was also continuously observed from samples sequenced from May to December 2020, during the first and second wave of SARS-CoV-2 infection, in the country. However, B, B.1, B.1.1, and B.1.1.1, which were present in the first wave, got the incidences of 1.2%, 4.82%, 7.23%, & 8.4% respectively in the second wave as well.

Among the new lineages of second wave the B.1.1.7, B.1.36.34 and C.23 had 12.1%, 4.8%, and 2.4% incidences respectively. The first case of B.1.36.34 was reported from Canada in May, 2020 and (four month later) in September 2020 the genomic sequence of this variant was reported from Pakistan [36]. It is most likely that both B.1.36.31, & B.1.36.34 were transmitted to Pakistan through the overseas Pakistani, whose significant number live in Canada & Australia and used to travel to Pakistan frequently. However, only 41 cases of B.1.36.34 are reported globally, till 10 July 2021, out of which 5 were reported from Pakistan (Table 2). The very small number of sequences of this lineage worldwide suggest its low transmissibility compared with other lineages. Though, B.1.1.7 (UK-VOC) had the high incidence in the second wave but it was reported almost at the end of the second wave and caused the highest infection rate in the third wave.

The B.1.36.17, B.1.36.24, B.1.1.25, B.1.1.372, B.1.1.413 were the minor lineages in second wave and had their first cases reported mainly from England and Wales [36] and are also likely to be transmitted to Pakistan from UK via Pakistani diaspora. Likewise, B.1.36.8, B.1.523, & AE.4 had their first case reported from India, Saudi Arabia, and Bahrain respectively in the early 2020 [37]. And they are also likely transmitted to Pakistan via international travelers.

### Third wave of SARS-CoV-2

The third wave of SARS-CoV-2 proved worst in the country with the highest number of infections (335,728) and deaths (7,849) compared with the previous waves. In the third wave, B.1.1.7, B.1.617.2 and B.1.351 were the major lineages for the SARS-CoV-2 based infections (Fig 11 & Fig 12). During this wave, first two cases of B.1.1.7 variants were reported on 20th December 2020 in the country and kept spreading continuously and got the highest incidence during the peak month of third wave. It is further evidenced by the submission of 108 (60.6%) genomic sequences of B.1.1.7 (UK VOC), only in the April 2021, out of the total 178 genomic sequence of this lineages from Pakistan, starting from 1^st^ December 2020 to 10 July 2021 (Fig. 4). The B.1.1.7 variant is estimated to have emerged in September 2020 and quickly become the dominant circulating variant in England[38]. In addition to Pakistan, this variant had been reported from 30 other countries.

The modeled trajectory of this variant in the Pakistan revealed rapid increase in its cases in early 2021, making it the predominant variant in the third wave (Fig 7, 8, & 9). Consistent, with our findings several other authors had reported that this variant had high transmissibility, virulence and death rates compared with other variants of SARS-CoV-2 [12, 39, 40]. Moreover, it is also worth mentioning that this variant had very high mutation rate (ranging from 45-50) (Fig. 7), in Pakistan, compared with other variants. Looking at the severity of the third wave, it is speculated that these mutations might have resulted in enhancing the transmissibility and pathogenicity of this variant in Pakistan.

B.1.617.2 lineage (Delta variant) was another variant which was observed almost at the end (May 2021) third wave of COVID-19 in Pakistan. It was originally reported from India in April 2020. Scientists have reported that Delta variant is 40-60% more contagious and virulent compared with B.1.1.7 [41, 42] and had caused a catastrophic wave of COVID-19 in India [43]. However, Pakistan remained lucky enough that this lineage was contained very shortly and in total only 24 genomic sequences of this variants had been reported from Pakistan, till 10-07-21. However, only this variant is known to have caused at least 4 million deaths in the world, mainly in India & UK [44].

B.1.351 lineage (Beta variant) was also reported during the third wave in Pakistan. Though this variant is known as South African variant but its first few cases were reported from England in the start of 2020 [37]. However, from Pakistan first case of this lineage was reported in May 2021 and it is highly likely that variant was transmitted to Pakistan from England through overseas Pakistanis. This variant is also known to have significantly greater transmissibility over alpha VOC [44-46]. However, only 30 genomic sequences of this variant are reported so far (10-07-2021) from Pakistan which are 12.9% of total genome sequences of SARS-CoV-2 submitted from Pakistan during the third wave of COVID-19. Our data revealed that B.1.1.7, B.1.351, and B.1.617.2 were the dominant lineages in the third wave, and this is the most likely reason for the severity of this wave in term of infections and deaths in the country.

## Materials and Methods

### Collection of the health data of COVID-19 patients

The data recorded on COVID-19 patients in different geographical regions of Pakistan as Diseased, Hospitalized, Deceased and Recovered were taken from official website of Pakistan (www.covid.gov.pk), for a period spanning from 3 January 2020 to 16 June 2021.

### Viral genome sequences

In total, 453 whole genome sequences of SARS-CoV-2, reported from Pakistan, were downloaded from GISAID (https://www.gisaid.org/) on 10 July 2021.

### Wave-wise categorization of SARS-CoV-2 genome sequences

The data were categorized into three waves on the basis of date of collection of samples. For the first wave which spanned from 1^st^ January 2020 to 31 October 2020, 89 whole genome sequences of SARS-CoV2 were available. Second wave ranged from 1^st^ November 2020 to 15^th^ February 2021 and for this wave 118 samples of SARS-CoV-2 genome sequences were submitted to GISAID. For third wave which spanned from Mid of February to the end of May 2021, a total of 246 whole genome sequences of SARS-CoV-2 were downloaded from the GISAID.

### Multiple Sequence Alignment of SARS-CoV-2 genomes

Sequence alignment of 453 Pakistani samples was performed using L-INS-I alignment method implemented in MAFFT (v7.480), by setting data type as nucleic acids with gap extend penalty of 0.123 and opening penalties default settings of 1.53 [47]. The sequences with more than 50 ambiguous bases were removed from the data file in order to minimize the number of false positive. This resulted in the removal of 53 samples (3, 35, & 15 samples from 1^st^, 2^nd^ & 3^rd^ wave respectively), and subsequently 400 samples were left for the analysis. The genome of SARS-CoV-2 reported from Wuhan, China, in December 2019, and available at GenBank with accession no. NC_045512.2 was used as a reference genome. Same method of alignment was used when sequences of each wave were aligned separately.

### SARS-CoV-2 Lineage Assignment

Phylogenetic Assignment of Named Global Outbreak Lineages (Pangolin) was used to describe the genomic lineages of the Pakistani SARS-CoV-2 sequences. The Pangolin tool follows the ‘Pango’ nomenclature system for classifying SARS-CoV-2 genomic sequences

### Construction of Phylogenetic tree with full length genomic sequences

To infer our SARS-CoV-2 time-scaled and divergence phylogenetic tree, selected global reference sequences were used for the Nextstrain analysis as of 10 July, 2021. This dataset consisted of 3,804 sequences sampled between 26th December 2019 to 10 July 2021 from Africa (541), Asia (937), Europe (727), North America (779), Oceania (512), and South America (566). A collection of 400 Pakistani SARS-CoV-2 sequences was added to generate an initial dataset of 4204 whole-genome sequences. Using the Nextstrain metadata [21] to identify the accessions of interest, the latest whole-genome sequence alignment from the GISAID database was downloaded. These 4204 whole genome sequences were aligned using MAFFT (v.7.480) [47], through multiple sequence alignment method, and manually edited by trimming the 5’ and 3’ untranslated regions and removing any gap only sites and low-quality sequence. The Wuhan/Hu-1/2019 (EPI_ISL_402125), sampled on December 31, 2019, from Wuhan, China was downloaded from the GISAID and used as reference genome. Finally, the maximum likelihood phylogenetic tree was built by using the Nextstrain pipelines which incorporates Augur for generation of phylogenetic tree and Auspice for visualizations. A coverage map was created using Nextclade (version 0.7.5), which performs banded Smith–Waterman alignment with an affine gap penalty.

### Wave-wise constrution of phylogenetic tree

Phylogenetic tree of each wave was inferred by using the Maximum Likelihood method based on amino acid substitution model of Jones Taylor Thornton (JTT) matrix-based model in Mega-X software. In each wave, initial phylogenetic tree(s) for the heuristic search were obtained automatically by applying Neighbor-Join and BioNJ algorithms to a matrix of pairwise distances estimated using the JTT model, and then finally, maximum likelihood phylogenetic tree of each wave was made by selecting the topology with superior log likelihood value.

## Data Availability

All the data used in this manuscript are publicly available from GISAID (https://www.gisaid.org/) and covid-19 website of Pakistan (https://covid.gov.pk/)

## Acknowledgement

We are thankful to all the laboratories that submitted their sequences to the GISAID platforms. We are also grateful to GISAID team for managing this platform.

